# Genetic determinants of cytokine production in activated human monocytes

**DOI:** 10.64898/2026.05.08.26352736

**Authors:** James J Gilchrist, Alexander J Mentzer, Luke Jostins, Seiko Makino, Vivek Naranbhai, Sara Danielli, Isar Nassiri, Julian C Knight, Benjamin P Fairfax

## Abstract

Monocyte function plays a central role in human health and mapping the genetic determinants of monocyte gene expression has provided insights into numerous disease processes. The relationship between genetic variation and functional cytokine secretion in response to immune stimuli remains poorly characterised however. To address this, we have quantified the production of 28 cytokines by monocytes from 366 healthy, European-ancestry donors following activation with lipopolysaccharide (LPS) and interferon gamma (IFNγ). By integrating these data with genomic and transcriptomic data from the same cells we robustly define the regulatory determinants of monocyte cytokine secretion. We identify four genome-wide significant loci affecting monocyte cytokine release, observing both *cis* and *trans* regulatory effects on cytokine release. These loci include multi-cytokine *trans* regulatory activity of the CCR5-Δ32 deletion on secretion of the CCR5-binding cytokines, MIP-1β and RANTES, and a *cis* regulator of PDGF-BB secretion, which colocalises with GWAS risk loci for ulcerative colitis and primary biliary cirrhosis. We further map the genetics of co-expression to establish relationships between RNA transcription and cytokine protein secretion. In doing so we identify marked enrichment of genes related to lipid metabolism in gene regulatory networks linked to cytokine secretion and identify that the COVID-19 risk locus at *OAS1* uncouples *OAS1* RNA expression from the secretion of 10 cytokines in response to LPS stimulation.

## Introduction

Monocytes are a heterogeneous group of innate immune cells. They are derived from the common myeloid progenitor and comprise ∼10% of blood leukocytes in the peripheral blood^1,2^. They form central mediators and effectors of the innate immune response, having both pro-inflammatory and regulatory functions, contributing to the clearance of pathogenic insults and the subsequent resolution of inflammation. Much of this functionality is mediated through the release of monocyte-derived cytokines, allowing the coordination of inflammatory responses across diverse immune cells. Monocytes and their cytokines thus have key roles in the pathogenesis of a broad range of human diseases, including infection, inflammatory disease, cardiovascular disease and cancer.

Genome-wide association studies (GWAS) have been highly successful in shaping our understanding of the genetics of inter-individual variability in disease risk^3^. Moreover, combining GWAS with functional genomic studies, for instance expression quantitative trait locus (eQTL) studies mapping the genetics of gene regulation, has been highly informative of human disease susceptibility, allowing the robust identification of disease-associated genes and the prioritisation of novel drug targets^4,5^. We have previously demonstrated that common genetic variation is a key determinant of gene expression^6–8^, splicing^9^ and epigenetic change^10^ in monocytes, and that regulatory variation is critically dependent on activation state. These context-dependent effects are highly informative of human phenotypic variation and disease. The effect of eQTL on human disease risk is likely to be largely mediated through protein abundance and function, however, large-scale mapping of protein QTL (pQTL) has demonstrated that eQTL and pQTL are commonly distinct^11^, highlighting the importance of mapping both the regulatory determinants of RNA expression of protein abundance. We know that, in the case of eQTL studies, mapping regulatory genetic determinants in bulk tissues, for instance in whole blood, will fail to detect many disease-relevant eQTL restricted to cell-type or activation state. It is likely that the same is true of pQTL, however, cell- and context-specificity have been relatively underexplored in pQTL studies. To address this, we mapped the human genetic and transcriptional determinants of secretion of 28 cytokines in primary human monocytes from 366 European-ancestry donors following *ex vivo* stimulation with either lipopolysaccharide (LPS) or interferon gamma (IFNγ). In doing so we describe the dynamics of monocyte cytokine production, defining its relationship to transcription, and identifying genetic determinants of monocyte cytokine production that are informative of human disease.

## Results

### Innate immune stimulation & cytokine secretion

We first sought to establish the trajectories of cytokine release following innate immune activation in monocytes and the extent to which these were shared and likely co-regulated. Following quality control, we compared monocyte secretion of 28 cytokines in 995 samples across four conditions: untreated (UT, n=78), 2 hours LPS (LPS2, n=263), 24 hours LPS (LPS24, n=314) and 24 hours IFNγ (IFNG24, n=340). LPS stimulation resulted in changes in the secretion of 19 of 28 cytokines tested (Fig. 1A, Table S1). There were two patterns to these changes, with 6 cytokines (IDO, IL-1β, IL-10, MIP-1α, MIP-1β, TNFα) showing early, sustained changes with significant perturbation at LPS2 and LPS24, and 13 cytokines (GROα, HGF, IL-1RA, IL-2, IL-21, IL-6, IP-10, LIF, MCP-1, PIGF-1, RANTES, SDF-1α, VEGF-A) showing isolated late changes with perturbation only seen at LPS24. The induction of 5 cytokines was noted at IFNG24: IDO, IL-18, IL-7, IP-10, TNFα. For all responsive cytokines, innate activity elicited an induction of secretion compared to unstimulated monocytes except for HGF, for which inhibition of secretion was noted at LPS24. Cytokine induction following LPS and IFNγ stimulation was highly coordinated, with hierarchical clustering of cytokine responses within each stimulation condition demonstrating distinct groups of co-regulated cytokines (Fig. 1E).

**Fig. 1:**
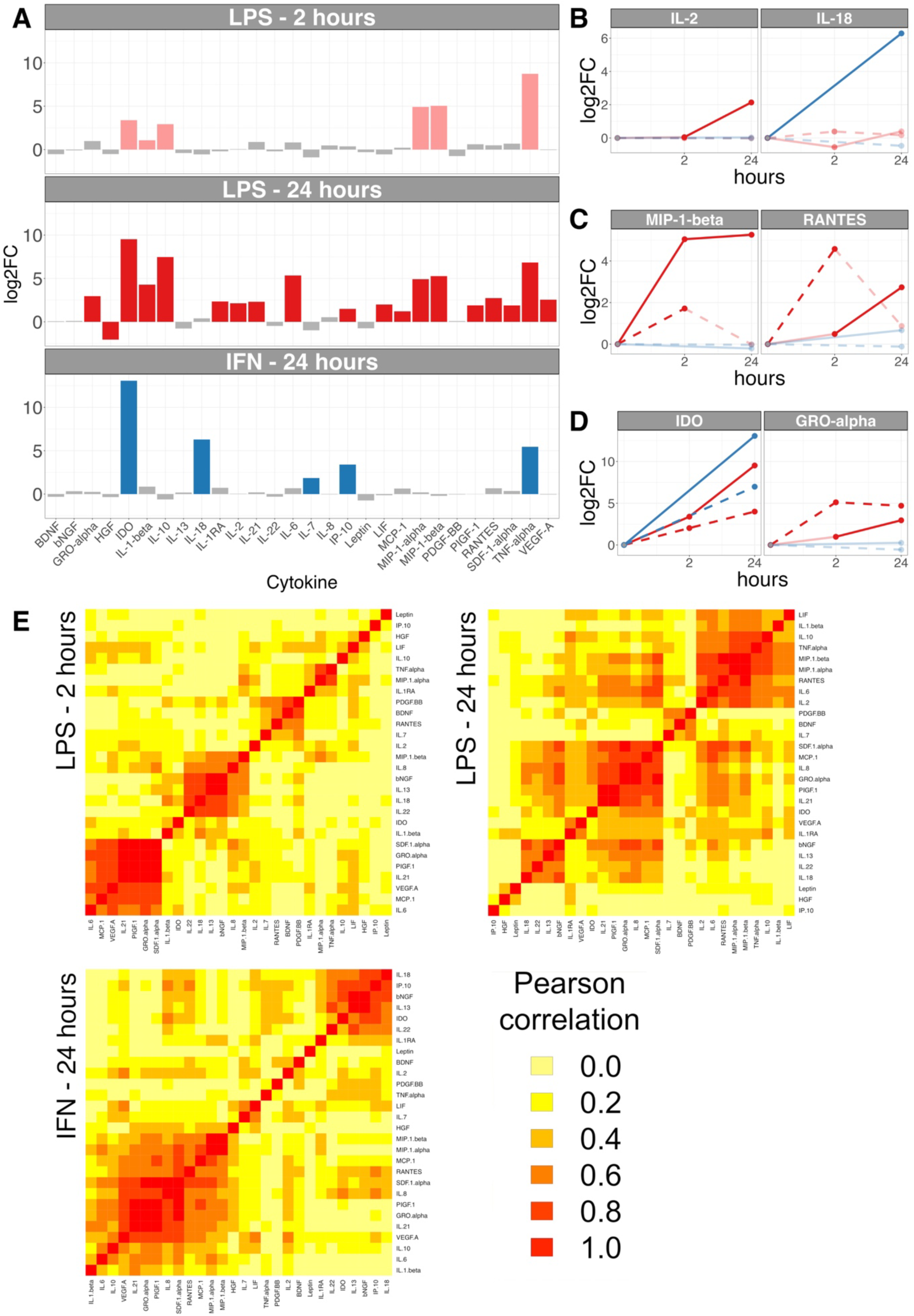
Monocyte cytokine secretion following innate immune stimulation. (**A**) Fold-change in cytokine secretion (n=28) following LPS stimulation for 2 and 24 hours and IFNγ stimulation for 24 hours. Cytokines with significant responses to innate stimulation (FDR<0.05, fold change ≥2) are coloured (LPS 2 hours, pink; LPS 24 hours, red; IFNγ, blue). log2FC, log2 fold change. (**B**) Examples of cytokines where secretion is significantly upregulated by innate stimulation with unchanged levels of RNA expression. (**C**) Examples of cytokines where secretion is significantly upregulated by LPS stimulation where RNA expression is transiently induced. For MIP1β (left) this results in a sustained increase in cytokine secretion. For RANTES (right) this results in increased secretion only at 24 hours. (**D**) Examples of cytokines where secretion is significantly upregulated by LPS stimulation with a sustained increased in RNA expression. For IDO (left) this results in a sustained increase in cytokine secretion. For GROα (right) this results in increased secretion only at 24 hours. Red lines represent responses to LPS, blue lines responses to IFNγ. Solid lines represent cytokine secretion, dashed lines represent RNA expression. Lines in bold represent significant changes (FDR<0.05, fold change ≥2). (**E**) Heatmaps depicting correlation of cytokine responses following LPS2, LPS24 and IFNγ stimulation. Colours represent pairwise absolute Pearson correlation coefficients, with cytokines ordered by hierarchical clustering.

Preceding or concurrent transcriptional responses of the respective genes were observed for 12 of 19 following LPS and 3 of 5 following IFNG24 (Fig. S1, Table S1). Similarly, significant transcriptional changes are typically followed by changes in cytokine secretion, with 15 of 22 changes in RNA expression being associated with changes in protein secretion. Changes in cytokine secretion are, however, seen in the absence of transcriptional changes for a subset of genes, with IL-2 secretion on LPS24 stimulation and IL-18 on IFNG24 stimulation occurring in the absence of significant increases in RNA expression at the time points assayed in our model (Fig. 1B). Following LPS stimulation, this pattern is only seen where cytokine changes are late, i.e. LPS24. Isolated transcriptional changes, without observed changes in cytokine secretion, are most frequently seen for downregulated transcripts, however both IL-8 and PDGF-BB exhibit increased RNA expression following LPS stimulation without significant changes in protein secretion. We observed two dominant patterns of transcriptional change resulting in altered cytokine secretion on LPS stimulation. Whereas in 5 of 12 cases, transcriptional changes were early and transient (Fig. 1C), 6 of 12 demonstrate sustained transcriptional changes across both 2 and 24 hour timepoints (Fig. 1D). Conversely, VEGFA specifically demonstrated only late induction of transcript and protein at LPS24, in keeping with the role of chronic, unresolved inflammation, promoting myeloid driven angiogenesis^12^.

### Cytokine RNA transcription as a predictor of secretion

We then explored the relationship between transcript and cytokine secretion across timepoints. In keeping with context-specific regulation of most cytokines, baseline transcript expression showed no relationship with magnitude of induced cytokine secretion (Fig. 2, Table S2) except for a very weak relationship between the cytokines RANTES and IP-10 and their respective transcripts, *CCL5* (r^2^=0.04) and *CXCL10* (r^2^=0.05), at LPS2. Interestingly, at LPS2 RANTES or IP-10 were not significantly induced, whereas at LPS24, when both cytokines are induced, baseline RNA expression no longer predicted secretion. Cytokine RNA expression following stimulation was more predictive of protein secretion, with a significant relationship for 39/84 RNA/cytokine pairs (FDR<0.05) at the same timepoint. Importantly, however, transcription in these cases typically explains a small proportion of the observed variance in cytokine secretion, with the maximal r^2^ of 0.56 noted for the RNA/cytokine pair: *CCL2*:MCP-1 (LPS2). These correlations are also highly timepoint dependent, for instance the RNA/cytokine correlation for *CCL2*:MCP-1 falls to r^2^=0.08 at LPS24, despite protein secretion increasing through to the later timepoint. This supports the importance of other processes such as translation, intracellular trafficking, decay and reabsorption, being further determinants of net cytokine secretion, their relative importance varying over time post-initial stimulation.

**Fig. 2.**
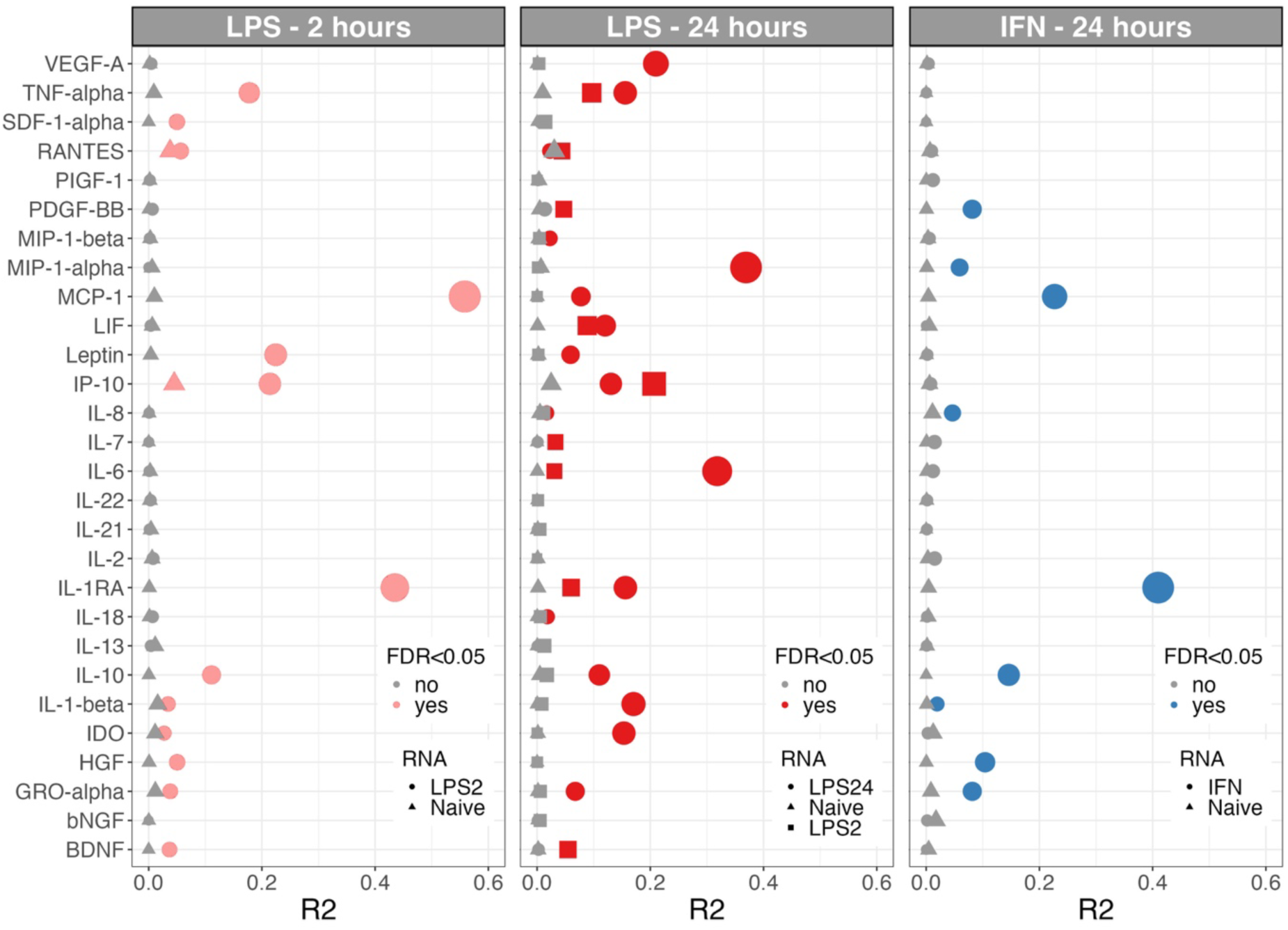
RNA transcription as a predictor of cytokine secretion in stimulated monocytes. Correlation between RNA transcription and protein secretion for each of 28 cytokines. Correlations are shown for protein secretion following LPS stimulation for 2 hours (left panel), 24 hours (middle panel) and IFNγ (right panel). Significant correlations are highlighted (pink/red/blue), and points are shaped according to the RNA timepoint protein secretion is correlated with (triangle, baseline; square, 2 hours; circle 24 hours). Point size is proportional to level of significance of the association. Correlation is calculated by linear regression. FDR, false discovery rate.

### Transcriptional state and cytokine secretion

To better define the dependency between transcriptional state and cytokine secretion, we analysed the relationship between baseline and stimulated monocyte transcriptomes and cytokine secretion. As for the relationship between cytokine RNA expression and protein secretion, we observed very limited association between baseline transcriptional state across all transcripts and cytokine secretion, with only 14 of 18,078 transcripts showing evidence of association (FDR<0.05, Table S3). Of note, *JUN* expression is significantly correlated with IL-1β secretion after IFNG24 (Fig. 3A, Pearson’s r=0.31, p=6.9×10^−9^). This association is specific to the baseline state, with no association between *JUN* and IL-1β at IFNG24 (p=0.78) or following LPS stimulation (LPS2 p=0.64, LPS24 p=0.9). IL-1β secretion is mediated by inflammasome activation, which converts inactive pro-IL-1β to its active form^13^. JUN encodes a constituent of the AP-1 TF, which regulates pro-IL-1β transcription^14^. Consistent with this, our data suggests a model whereby baseline *JUN*, and thus AP-1 activity, determines early transcription and translation of pro-IL-1β, which is then primed for activation and secretion following IFN-γ-induced inflammasome activation, and that monocytes with JUN-high transcriptional states may be primary drivers of IL-1β secretion. Secondly, across all 3 stimulation conditions, BDNF and PDGF-BB secretion are significantly correlated with the baseline expression of a group of 13 genes (Table S3, Fig. 3B), including *CAVIN2*, *TUBB1*, *GP9* and *PPBP*. Those 13 genes are highly enriched for platelet-specific genes (p=5.2×10^−9^, fold-change=17.6). This suggests that this cytokine-associated RNA expression signature is a marker of the proportion of monocyte-platelet aggregates (MPA). In keeping with this, BDNF, a neurotrophin family growth factor, is stored^15^ and released^16^ by platelets and MPAs are a major source of BDNF in stimulated PBMC cultures^17^. Similarly, PDGF-BB is secreted by platelets, inducing chemotaxis in fibroblasts, smooth muscle cells and monocytes, but is also synthesised and secreted by stimulated monocytes and macrophages in the absence of platelets^18^. Secretion of BDNF or PDGF-BB is stable across monocyte stimulation conditions (Fig. S1), and the effect of the baseline platelet signature on secretion of both cytokines is observed in UT, LPS2, LPS24 and IFNG24 monocytes. This suggests a model whereby BDNF or PDGF-BB are either directly platelet-derived or are monocyte-derived in a manner licenced by platelet presence^19^.

**Figure 3.**
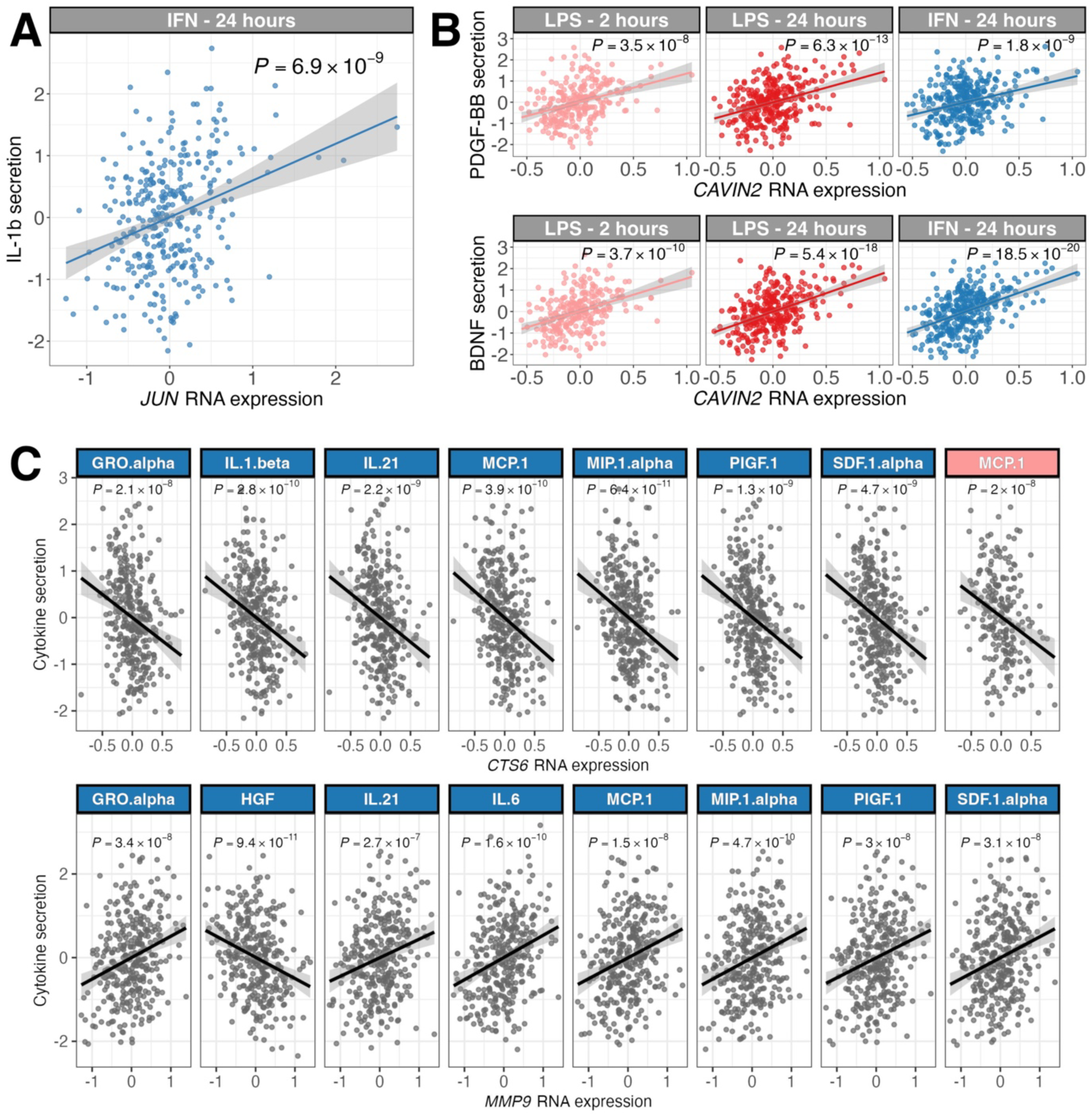
Genome-wide transcriptional predictors of cytokine secretion. (**A**) Correlation between baseline *JUN* transcription and IL-1β secretion following 24 hours of IFNγ stimulation. (**B**) Correlation between *CAVIN2* transcription at baseline and BNDF/PDGF-BB secretion in stimulated monocytes (LPS2, LPS24, and IFN24). (**C**) Correlation between MMP9 transcription in stimulated monocytes and the secretion of cytokines. Plots highlighted with blue facets depict RNA transcription and cytokine secretion in IFNγ-stimulated monocytes, with plots highlighted with pink facets depicting monocytes stimulated with LPS for 2 hours.

The stimulated transcriptional state is markedly more associated with cytokine secretion across all 3 stimulation conditions (Table S4). At LPS2, RNA expression is associated with cytokine secretion for 32 gene:cytokine pairs (FDR<0.05), including 8 distinct cytokines. The number of correlated RNA:cytokine pairs at LPS24 is similar: 27 pairs including 7 distinct cytokines. We observed more marked correlation between the transcriptome and cytokine secretion following in the IFNG24 dataset, with 148 correlated RNA:cytokine pairs for 11 cytokines. In keeping with the co-regulation of cytokine responses (Fig. 1E), we observe several instances of shared correlated transcription across cytokines. For instance, expression of both *MMP9*, encoding matrix metalloproteinase-9, and *CST6*, encoding cystatin-M, is each associated with induction of a group of 8 overlapping cytokines, including GROα, IL-21, MCP-1, MIP-1α, PIGF-1and SDF-1α (Fig. 3C). The largest transcriptional network associated with secretion of a single cytokine was a 43 gene network associated with MCP1 secretion following IFNG24 stimulation. This network is highly enriched for mediators of the unfolded protein response (UPR, R-HSA-381119, p=5.2×10^−8^, fold-change=24.4), as well as IRE1α (R-HSA-381070, p=1.8×10^−7^, fold-change=31) and XBP1 (R-HSA-381038, p=1.4×10^−7^, fold-change=32.2) mediated UPR signalling. IRE1α acts as a sensor of endoplasmic reticulum stress, triggering the UPR through splicing of XBP1. XBP1 in turn increases ER capacity, through chaperone transcription/translation and increase of ER size, and induction of the secretory pathway^20^. Moreover, innate immune stimulation activates XBP1 via IRE1α signalling in macrophages and drives pro-inflammatory cytokine induction independent of ER stress^21^.

### Genetic determinants of cytokine secretion

Following quality control, we included 5,326,275 SNPs in a GWAS of cytokine secretion across the induced conditions (LPS2, n=263; LPS24, n=314; IFNG24, n=340). Given the observed co-correlation in cytokine responses (Fig. 1E), we performed genome-wide association analysis using multivariate tests (MANOVA) to reflect the correlated nature of the outcomes and to maximise study power, as well as univariate GWAS for each of the 28 cytokines passing QC. We considered *P*<1.7×10^−8^ to be significant for the MANOVA GWAS and *P*<6.0×10^−10^ for the univariate GWAS. Inspection of QQ plots (Fig. S2) and genomic inflation factors (maximum λ=1.02, Table S5) did not indicate uncontrolled confounding variation. We identified 4 genetic loci (2 *cis*, 2 *trans*) significantly associated with cytokine secretion at experiment-wide significance: 3 in the MANOVA GWAS and 1 locus in the univariate analysis (Table 1, Fig. 4A, B, C).

**Figure 4.**
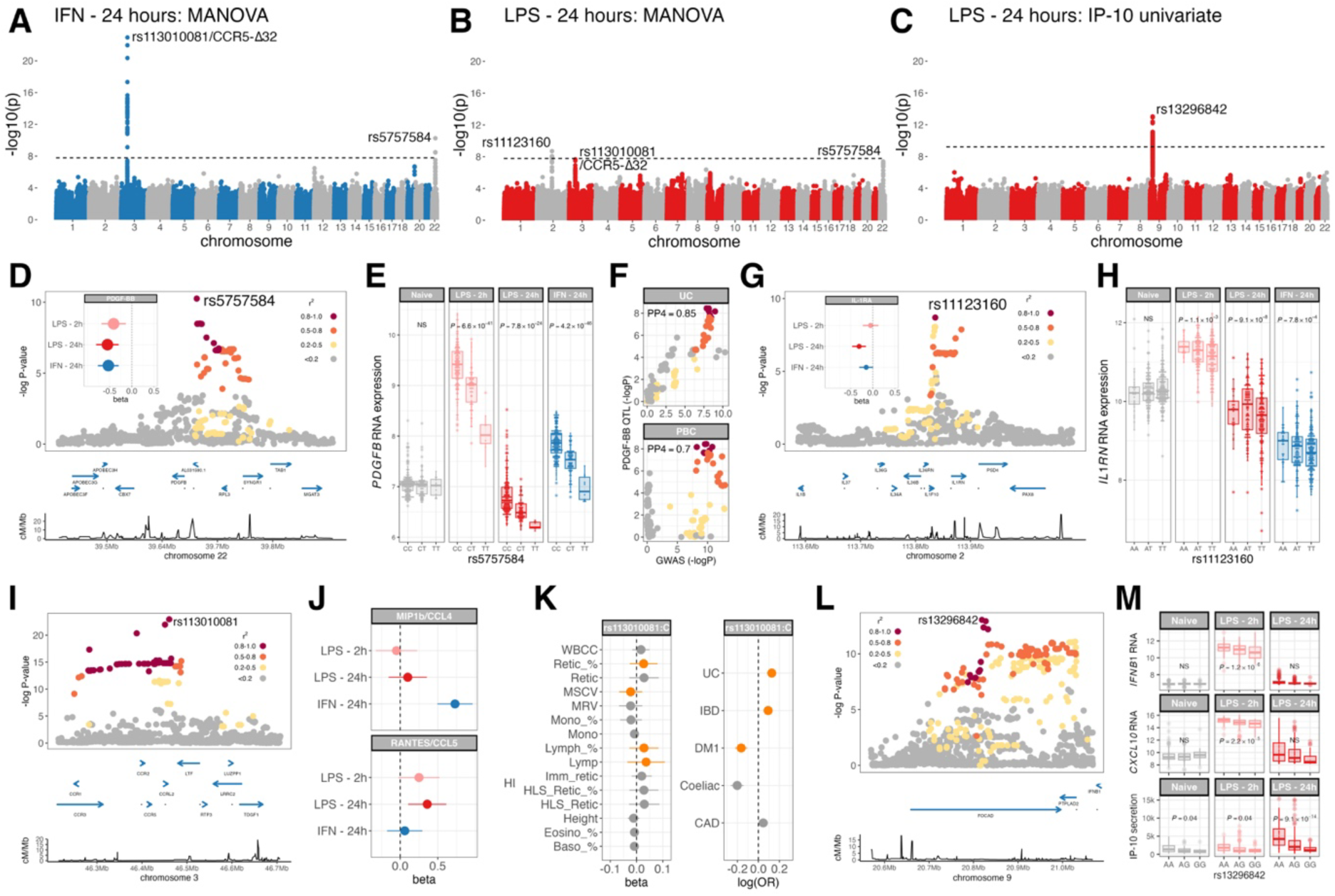
Genome-wide association study of cytokine secretion in stimulated monocytes. Manhattan plots for multivariate genome-wide association analysis (MANOVA) of cytokine (n=28) secretion from monocytes stimulated for 24 hours with IFNγ (**A**) and LPS (**B**). (**C**) Manhattan plot of univariate genome-wide association analysis of IP-10 secretion from monocytes stimulated with LPS for 24 hours. Dashed lines represent experiment-wide significance thresholds: *P*<1.7×10^−8^ (multivariate), *P*<6.0×10^−10^ (univariate). Significant associations are labelled. (**D**) Regional association plot of a multivariate association test (MANOVA) of cytokine (n=28) secretion in IFNγ-stimulated monocytes at the *PDGFB* locus. Points are coloured according to degree of linkage disequilibrium (r^2^) to the lead SNP (rs5757584). The inlaid forest plot depicts the univariate association between rs5757584:T allele carriage and PDGF-BB secretion in stimulated monocytes. (**E**) Box and whisker plots depicting the association between rs5757584 genotype and *PDGFB* RNA expression in naïve and stimulated monocytes. P-values are calculated by linear regression. (**F**) Evidence for colocalization between genetic signals of ulcerative colitis (UC) and primary biliary cirrhosis (PBC) risk and the PGGF-BB cytokine secretion QTL in IFNγ-stimulated monocytes. Points are coloured according to degree of linkage disequilibrium (r^2^) to the lead SNP (rs5757584). (**G**) Regional association plot of a multivariate association test (MANOVA) of cytokine (n=28) secretion in LPS-stimulated (24 hours) monocytes at the *IL1RN* locus. Points are coloured according to degree of linkage disequilibrium (r^2^) to the lead SNP (rs11123160). The inlaid forest plot depicts the univariate association between rs11123160:T allele carriage and IL-1RA secretion in stimulated monocytes. (**H**) Box and whisker plots depicting the association between rs11123160 genotype and *IL1RN* RNA expression in naïve and stimulated monocytes. P-values are calculated by linear regression. (**I**) Regional association plot of a multivariate association test (MANOVA) of cytokine (n=28) secretion in IFNγ-stimulated monocytes at the *CCR5* locus. Points are coloured according to degree of linkage disequilibrium (r^2^) to the lead SNP (rs113010081). (**J**) Forest plots describing the univariate association between MIP-1β (top) and RANTES (bottom) secretion and rs113010081/CCR5-Δ32 in stimulated monocytes. (**K**) Effect of rs113010081:C/CCR5-Δ32 on continuous phenotypic traits in the UK Biobank (left) and human disease risk (right). Diseases/traits for which there is evidence of colocalization with rs113010081/CCR5-Δ32 are highlighted (orange). (**L**) Regional association plot of a univariate association test of IP-10 secretion in LPS-stimulated (24 hours) monocytes at the *IFNB1* locus. Points are coloured according to degree of linkage disequilibrium (r^2^) to the lead SNP (rs13296842). (**M**) Box and whisker plots depicting the association between rs13296842 genotype and *IFNB1* RNA expression (top), *CXCL10* (encodes IP-10) RNA expression (middle) and IP-10 protein secretion in naïve and LPS-stimulated monocytes. P-values are calculated by linear regression. Box and whisker plots; boxes depict the upper and lower quartiles of the data, and whiskers depict the range of the data excluding outliers (outliers are defined as data-points >1.5X the inter-quartile range from the upper or lower quartiles).

We identify a *cis*-acting locus to *PDGFB*, encoding PDGF-BB, located at chr22:39,563,208-39,761,255, 22kb upstream from the *PDGFB* transcription start site, comprising 31 SNPs in the 95% confidence interval, with rs5757584:T (chr22:39,662,550:C:T) the peak expression associated SNP. Post IFNG24, rs5757584 associated with cytokine secretion at experiment-wide significance (MANOVA F=4.3, p=5.6×10^−11^), with additional evidence of association with cytokine secretion following the LPS24 stimulation (MANOVA F=2.8, p=9.5×10^−6^). These associations are driven by reduced PDGF-BB secretion in both treatments (IFNG24, β=-0.53, p=6.8×10^−9^, LPS24 β=-0.55, p=1.5×10^−8^) in rs5757584:T carriers (Table S6). There is also evidence of a concordant effect of rs5757584:T genotype on PDGF-BB secretion in the LPS2 stimulation (β=-0.41, p=1.2×10^−4^).

Analysis of *PDGFB* transcription demonstrated association with rs5757584 allele in IFNG24 (β=-0.4, p=4.2×10^−46^) and LPS-stimulated (LPS2, β=-0.61, p=7.6×10^−40^; LPS24, β=-0.27, p=7.8×10^−24^) monocytes, but not in unstimulated monocytes (β=-0.01, p=0.58), indicating rs5757584 association with PDGF-BB secretion is mediated via stimulus induced transcription. Consistent with these regulatory effects, rs5757584 overlaps an enhancer active in monocytes (ENSR22_9KKRT, chr22:39,662,400-39,662,858) and with TF binding sites in lymphoblastoid cell lines, including NF-κB binding sites in TNF-treated cells (Table S7). Interestingly, however, while *PDGFB* transcript at LPS2 is predictive of protein secretion at LPS24, RNA expression at 24 hours is not (Fig. 2). This suggests that while their genetic predictors are shared, it is variation in early transcriptional responses that is the critical determinant of PDGF-BB secretion. PDGF-BB secretion correlates with abundance of platelet-specific transcripts (Fig. 3B), potentially marking increased MPA numbers. It is possible that increased PDGF-BB secretion could represent the consequence of greater numbers of MPAs, or that increased secretion could be a driver of MPA formation. To test this, we assessed the effect of rs5757584 genotype on the RNA expression of platelet-specific transcripts in naïve monocytes, observing no significant effect of genotype either on platelet-specific transcription globally (permutation p=0.59) or on the expression of PDGF-BB associated transcripts specifically (n=7, p>0.05). In keeping with this, the effect of rs5757584 genotype on PDGF-BB secretion is independent of the effect of the baseline transcriptional signature, with the effect of rs5757584 genotype on cytokine secretion unaltered by conditioning on baseline *CAVIN2* expression. This suggests that platelet presence and PDGFB transcription are important, distinct determinants of PDGF-BB secretion. The genetic determinants of PDGF-BB secretion we identify here are informative of human health and disease. Variation determining PDGF-BB secretion colocalise with genetic risk loci for ulcerative colitis (PP4=0.85) and to a lesser extent primary biliary cirrhosis (PP4=0.70, Table S8).

The second *cis*-acting locus mapped to *IL1RN,* encoding IL1-RA, comprises 198 SNPs (95% CI) at chr2:113,739,532-113,897,953, with the peak association at rs11123160, chr2:113,836,165:A:T (Fig. 4G). This was associated with cytokine secretion in the LPS24 dataset (MANOVA F=3.9, p=2.0×10^−9^), but not after LPS2 (MANOVA F=1.27, p=0.17) or IFNG24 (MANOVA F=1.36, p=0.11). The effect of rs11123160 genotype on cytokine secretion is largely driven by its effect on IL-1RA secretion (24 hours LPS, β=-0.31, p=6.5×10^−5^, Table S9). Consistent with this, rs11123160 is an eQTL for *IL1RN* transcription in stimulated monocytes (LPS24, β=-0.22, p=9.1×10^−8^), and overlaps a monocyte-active enhancer 27kb upstream of *IL1RN* (ENSR2_CF7XW, chr2:113,835,428-113,837,397) and TF binding sites including for STAT1 in IFNα-treated K562 cells (Table S10). There is evidence of shared effects between the determinants of RNA transcription and protein secretion in the LPS24 dataset (PP4=0.79). Consistent with a shared genetic determinant, *IL1RN* transcript is highly predictive of IL-1RA protein secretion in each stimulation condition (Fig. 2). There is no strong evidence for colocalization between the genetic determinants of IL-1RA secretion and human phenotypic traits/disease (Table S11).

In *trans*, rs113010081 (chr3:46,457,412:T:C) is highly predictive of cytokine secretion in both LPS24 (MANOVA F=3.6, p=2.2×10^−8^) and IFNG24 (MANOVA F=8.1, p=1.2×10^−23^) datasets. This variant is in high LD (r^2^=0.977) with the CCR5-Δ32 deletion in the UK Biobank population, with the rs113010081:C allele tagging the CCR5-Δ32 frameshift mutation^22^ that confers resistance to human immunodeficiency virus (HIV) infection by introducing a non-functional receptor^23^. The CCR5 receptor binds the chemokines MIP-1β (encoded by *CCL4*) and RANTES (encoded by *CCL5*), and the secretion of both chemokines is affected by rs113010081 genotype in a stimulation-specific manner (Table S12). MIP-1β secretion is significantly increased post IFNG24 according to rs113010081:C/CCR5-Δ32 carriage (β=0.73, p=1.2×10^−9^), with no effect on RANTES secretion (β=0.06, p=0.61). However, post LPS, RANTES secretion is increased by rs113010081:C/CCR5-Δ32 carriage, most significantly following 24 hours of stimulation (LPS24 β=0.36, p=4.9×10^−3^), with a weak effect at 2 hours (LPS2 β=0.25, p=0.07). A nonfunctional CCR5 receptor would ablate any negative feedback the receptor confers for MIP-1β and RANTES release, which would tally with our observations. However, rs113010081/CCR5-Δ32 has no detectable effect on CCL4/CCL5 RNA expression in naïve or stimulated monocytes (Fig. S3), suggesting that the CCR5-Δ32 effect on cytokine concentration maybe independent of transcription. Additional to the association with HIV resistance, rs113010081/CCR5-Δ32 is linked to divergent phenotypic traits and disease risk including blood count indices, reduced risk of inflammatory bowel disease (IBD) and type 1 diabetes (Fig. 4K, Table S13).

The second *trans* locus was to rs13296842 (chr9:20,818,519:A:G), post LPS24, which associated with IP-10 secretion (LPS24 β=-0.56, p=9.1×10^−14^, Fig. 4L), with only a marginal effect on IP-10 secretion observed in LPS2 (β=-0.17, p=0.04, Fig. 4M) (Table S14). Notably, this locus forms a *trans* master regulator of LPS-induced transcription, mediated through an *IFNB1 cis* eQTL^7^ after 2h LPS, with downstream *trans* effects noted at 24h^7,24^. In keeping with this, rs13296842 genotype is associated with *IFNB1* expression in *cis* and *CXCL10* RNA (encoding IP-10) expression in *trans* post LPS (Fig. 4M). The kinetics of this relationship being akin to that observed with PDGF-BB, with transcript expression being observed early (after 2 hours of stimulation) and effects on protein expression being largely observed later.

Importantly, the genetic effects that we observe here on cytokine secretion are frequently distinct from those seen in large-scale experiments to map pQTL in whole blood^25^. For instance, while the effect of CCR5-Δ32 on MIP-1β secretion in IFNG24-stimulated monocytes is seen in whole blood without activation (UK Biobank, β=0.6, p=1×10^−625^), the effect of CCR5-Δ32 on RANTES stimulation following LPS stimulation is not (β=-0.005, p=0.69). Similarly, the effect of rs11123160 on IL-1RA secretion in LPS-stimulated monocytes is also seen as a significant pQTL in whole blood (UK Biobank, β=-0.17, p=4.1×10^−89^), while the effects of rs5757584 and rs13296842 on stimulated PDGF-BB and IP-10 secretion are not (UK Biobank, β=0.02, p=0.07; β=-0.005, p=0.47).

### Genetic determinants of the relationship between transcription and cytokine secretion

Polymorphisms that modify the expression relationships between genes can form co-expression QTL (coExQTL)^26,27^ that are highly informative of gene regulatory networks. Specifically, these *cis* eQTL disrupt the correlation structure between genes, potentially coordinated by a specific TF. Co-expression QTL mapping experiments have focussed on *cis* genetic variation modulating the co-expression relationship between pairs of RNA transcripts. However, induced cytokine secretion is highly co-regulated (Fig. 1E), and, as demonstrated, co-ordinated with transcript levels (Fig. 3). To assess for coExQTL that modify relationships between RNA transcription and cytokine secretion we performed coExQTL mapping to *cis* eQTL displaying significant effects on gene expression (FDR<1×10^−5^), identifying 1,756, 2,315 and 2,778 significant *cis* determinants of gene expression in LPS2, LPS24 and IFNG24 datasets respectively (Tables S15-17). We then correlated *cis* gene expression for each eQTL with secretion of cytokines under each stimulation condition, testing if these correlation relationships were modified by *cis* eQTL genotype.

We identified coExQTL for 239 RNA-cytokine secretion pairs in stimulated monocytes at experiment-wide significance (FDR<0.05), comprising 88 *cis* eQTL interacting with the secretion of 25 cytokines. The majority of these were observed in LPS24 (n=216), with 8 identified in LPS2 and 18 in IFNG24 (Tables S18-20). Of the 88 *cis* eSNPs, 12 overlap with TF binding sites in myeloid (K562) cells, and 40 are informative of human phenotypic variation or disease risk. In keeping with the observed co-regulation of cytokine secretion (Fig. 1E), we identify several instances of coExQTL involving transcription of a single gene and multiple secreted cytokines. For instance, rs7270354 (chr20:44,607,661:G:A) is a cis eQTL for *PLTP* in IFNG24 samples, and genotype at rs7270354 acts as a coExQTL between *PLTP* transcription and the secretion of 6 cytokines with IFNG24: GROα, IL-10, IL-21, IL-6, PIGF-1 and VEGF-A (Figure 5A, Table S20). *PLTP* encodes a lipid transfer protein with multifaceted functions, that has been shown to attenuate LPS responses^28^. Genetic variation at rs7270354 is informative of coronary artery disease risk (Table S20), with the disease risk allele (A) being associated with increased *PLTP* RNA expression in stimulated monocytes and enhanced positive association between *PLTP* RNA expression and IFNG24 cytokine secretion (Fig. 5A). Similarly, rs6145, an LPS24 context-specific eQTL for *VLDLR*, is a coExQTL for *VLDLR* expression and secretion of 5 cytokines at LPS24: TNF, IL-1β, IL-10, MIP-1α and MIP-1β. *VLDLR* encodes the very-low density lipoprotein receptor which, in tandem with *PLTP* coExQTL, suggests a central role for lipoprotein metabolism within the gene regulatory networks controlling monocyte cytokine responses. Supporting this, genes with coExQTL with cytokine secretion are enriched (Fig. 5C, Table S21 & S22) for lipid metabolic processes (GO:0006629, fold-change=3.7, p=0.008), familial hyperlipidemia (DOID:1168, fold-change=8.2, p=7.7×10^−5^) and lipid metabolism disorders (DOID:3146, fold-change=7.3, p=1.4×10^−4^).

**Figure 5.**
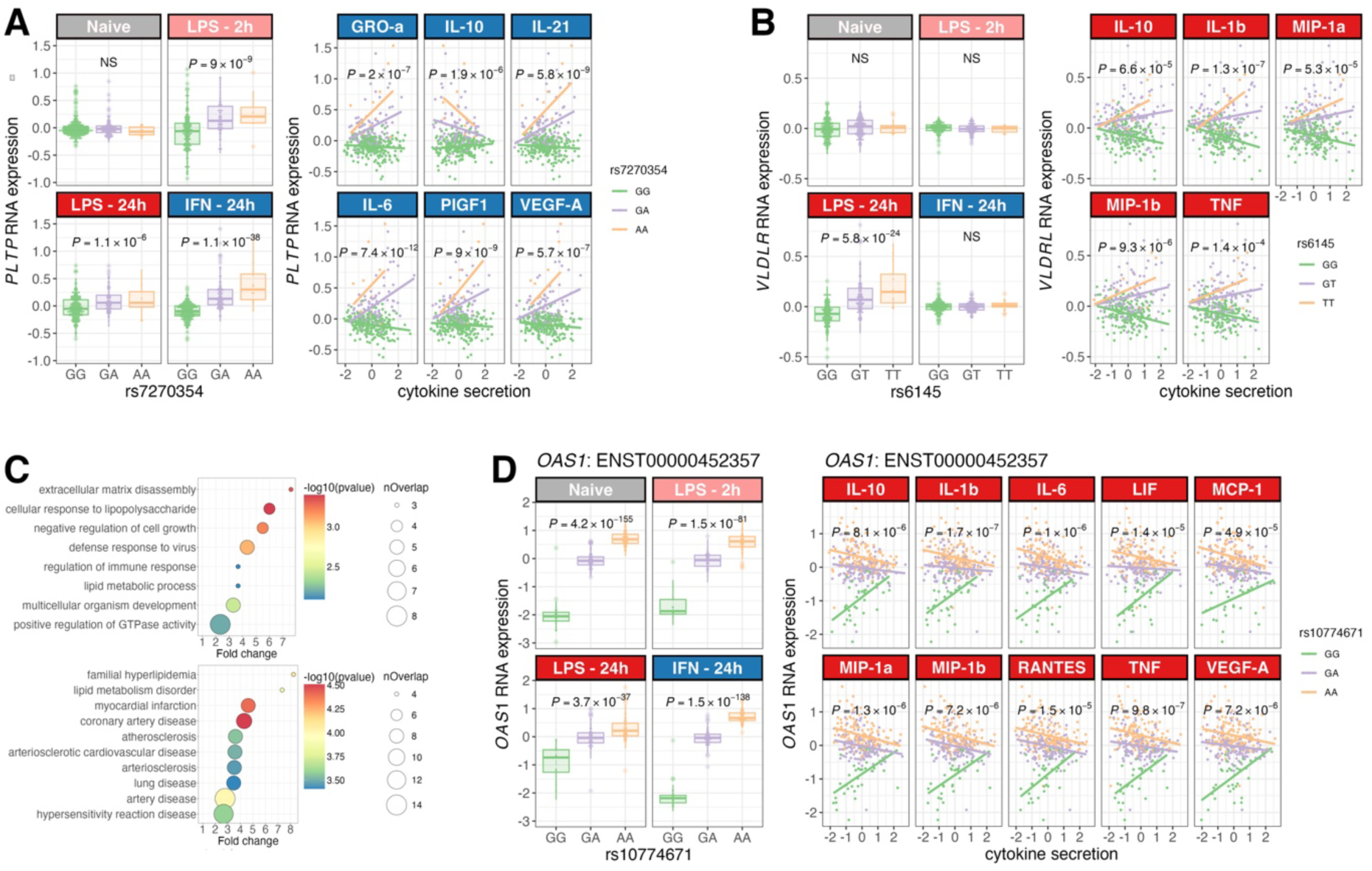
Genetics of the relationship between RNA expression in monocytes and cytokine secretion. (**A**) Box and whisker plots depicting the association between rs7270354 genotype and *PLTP* RNA expression in naïve and stimulated monocytes (left). Evidence for rs7270354 acting as a co-expression QTL between *PLTP* RNA expression and cytokine secretion (GRO-α, IL-10, IL-21, IL-6, PIGF1, VEGF-A) in IFNγ-stimulated monocytes (right). Points are coloured according to rs7270354 genotype, with linear regression lines describing the per-genotype relationship between *PLTP* RNA expression and cytokine secretion. P-values represent interaction terms in a linear regression model between genotype and cytokine secretion. (**B**) Box and whisker plots depicting the association between rs6145 genotype and *VLDLR* RNA expression in naïve and stimulated monocytes (left). Evidence for rs6145 acting as a co-expression QTL between *VLDLR* RNA expression and cytokine secretion (IL-10, IL-1β, MIP-1α, MIP-1β, TNF) in LPS-stimulated (24 hours) monocytes (right). Points are coloured according to rs6145 genotype, with linear regression lines describing the per-genotype relationship between *VLDLR* RNA expression and cytokine secretion. P-values represent interaction terms in a linear regression model between genotype and cytokine secretion. (**C**) Pathway enrichment analysis using Gene Ontology Biological Processes (top) and Disease Ontology (bottom) ontologies. Points are coloured according level of significance of enrichment among genes with coExQTL for cytokine secretion and sized according to the number of genes overlapping with a given pathway set. (**D**) Box and whisker plots depicting the association between rs10774671 genotype and *OAS1* ENST00000452357 RNA expression in naïve and stimulated monocytes (left). Evidence for rs10774671 acting as a co-expression QTL between *OAS1* ENST00000452357 RNA expression and cytokine secretion (IL-10, IL-1β, IL-6, LIF, MCP-1, MIP-1α, MIP-1β, RANTES, TNF, VEGF-A) in LPS-stimulated (24 hours) monocytes (right). Points are coloured according to rs10774671 genotype, with linear regression lines describing the per-genotype relationship between *OAS1* ENST00000452357 RNA expression and cytokine secretion. P-values represent interaction terms in a linear regression model between genotype and cytokine secretion. Box and whisker plots; boxes depict the upper and lower quartiles of the data, and whiskers depict the range of the data excluding outliers (outliers are defined as data-points >1.5X the inter-quartile range from the upper or lower quartiles).

Lastly, we identify a coExQTL at rs10774671 (chr12:112,919,388:G:T) for *OAS1* RNA transcript and secretion of 10 cytokines following LPS24 stimulation: IL-10, IL-1β, IL-6, LIF, MCP-1, MIP-1α, MIP-1β, RANTES, TNF and VEGF-A (Fig. 5D). This SNP forms a splice acceptor variant at the 5’ end of the 5^th^ intron of the ENST00000202917 *OAS1* transcript and is a risk locus for severe COVID-19^29^. We have previously demonstrated variation in high linkage disequilibrium with rs10774671 (rs10735079, 1000GP GBR population r^2^=0.86, D’=0.93), is a splice QTL (sQTL) for *OAS1* in LPS and IFNγ stimulated monocytes^9^, directing *OAS1* isoform usage. Carriage of the rs10735079:A allele (proxy of the rs10774671:A allele) is associated with increased expression of the ENST00000452357 *OAS1* isoform and decreased expression of the ENST00000202917 isoform, an effect reproducible using isoform-specific probes within our array data (Fig. S4). The effect we observe of rs10774671 as a coExQTL on cytokine secretion appears to be restricted to the ENST00000452357 isoform, with no significant effect observed for either the canonical *OAS1* isoform (ENST00000445409) or ENST00000202917 (Fig. S5). This transcript-specific effect is consistent with our previous observation that coExQTL governing RNA regulatory networks are frequently observed at transcript rather than gene resolution^9^. Importantly, these co-expression effects are not observable at the RNA level, with rs10735079 having no significant coExQTL effect between *OAS1* RNA and cytokine RNA expression (Fig. S6).

## Discussion

By combining analysis of multiplexed secreted cytokine protein abundance with cellular transcriptomics and genotyping, in a well-characterised model of monocyte innate immune activation in a large cohort of individuals, we provide new insights into the drivers, regulation and dynamics of cytokine production in human immune cells. We associate a range of phenotypic and disease risk loci with induced cytokine responses, of which a subset involve eQTL and regulatory networks. Notably, these determinants are often distinct from those observable in steady-state biobank-scale studies defining the circulating plasma proteome in health, emphasising the dissimilarity between baseline and induced cytokine responses.

LPS stimulation is associated with increased abundance of a broader set of secreted cytokines compared to IFNγ stimulation, a finding in keeping with the broader effects of LPS on monocyte DNA methylation^10^, but contrasting with the comparable transcriptional changes we observe induced by LPS and IFNγ^7^. While cytokine induction commonly occurs in the context of preceding transcriptional changes, this is not universal and late cytokine induction is frequently seen without concurrent or preceding transcriptional changes. This may reflect the limited temporal coverage of our transcriptional analysis, missing the relevant timepoint for induced RNA response. However, even when cytokine secretion followed transcriptional changes, transcription was only seen to explain a modest proportion of the variability in cytokine secretion, indicating key regulatory effects at levels including translation, trafficking and secretion. These observations are similar to T cell cytokine responses which are in part determined by the translation of pre-formed cytokine mRNA, as opposed to induced transcription^30,31^.

Although we found baseline transcriptional state beyond the specific cytokine showed limited prediction of cytokine secretion, informative exceptions were observed. For example, a baseline transcriptional signature enriched for platelet-expressed genes, which we hypothesise correlated with MPAs among cultured monocytes, is predictive of secretion of PDGF-BB and BDNF across stimulation conditions. Complementary to this, we also identify a proximal genetic determinant of PDFG-BB secretion, rs5757584, which functions as both an eQTL for *PDFGB* RNA expression and a QTL for PDFG-BB secretion in stimulated monocytes. These determinants are independent of each other, suggesting MPA presence may be enhancing PDGF-BB secretion independent of transcription changes, either through translation and secretion of PDGF-BB from associated platelets, or through licensing of PDGF-BB secretion from monocytes within MPAs. MPA formation has been previously shown to alter monocyte phenotype^32^, upregulating CD16 expression, and have been observed at increased frequencies in cardiovascular disease^33^, severe COVID-19^34^ and autoimmune diseases, including rheumatoid arthritis and systemic lupus erythematosus^35^. In keeping with the potential role for MPAs in autoimmune disease pathogenesis, rs5757584:T is associated with ulcerative colitis and primary biliary cirrhosis risk, with decreased *PDGFB* transcription and cytokine production associating with increased risk of autoimmune disease.

Genetic determinants of cytokine production distinct from those governing gene expression changes were noted at CCR5-Δ32, proxied by rs113010081, which is a determinant of secretion MIP-1β (encoded by *CCL4*) and RANTES (encoded by *CCL5*), for which CCR5 is the receptor. CCR5-Δ32 is a frameshift mutation resulting in the expression of a non-functional CCR5 molecule, the expression of which associated with increased cytokine production in stimulated monocytes. This effect operates independently of transcription of either *CCL4* or *CCL5* and may reflect an anti-inflammatory role of CCR5 via scavenging proinflammatory cytokines and chemokines at the point of resolution of acute inflammation^36^. Carriage of CCR5-Δ32 is associated with increased risk of IBD, specifically ulcerative colitis, suggesting that loss of CCR5 as a constraint on inflammation may contribute to IBD risk.

Lastly, we leverage genotype, transcriptome and cytokine secretion data to identify RNA:protein coExQTL, revealing relationships between gene regulatory networks and cytokine secretion. We identify that coExQTL genes linked to cytokine secretion are enriched for genes with roles in lipid metabolism. For example, a coExQTL at *PLTP* (encodes phospholipid transfer protein) is linked to secretion of a group of 6 cytokines at IFNG24, and a coExQTL at *VLDLR* (encodes very-low density lipoprotein receptor) is linked to secretion of a group of 5 cytokines in LPS-stimulated cells. This observation complements a growing appreciation of the role of lipid and lipoprotein metabolism in the regulation of inflammation, further linking atherosclerosis to innate immune responses. For instance, genetic variation at *VLDLR* has been previously linked to the regulation of steady-state cytokine levels in health^37^. In addition, PLTP itself can directly transfer LPS from bacteria to lipoproteins, acting to attenuate inflammatory responses^28^, and PLTP activity increases in patients with sepsis^38^. Indeed, marked perturbations in circulating lipid profiles are a reproducible feature of adult and paediatric sepsis^39^. We also identify coExQTL at *OAS1*, which disrupt the relationship between expression of the *OAS1* ENST00000452357 isoform and secretion of a group of 10 cytokines in response to LPS stimulation. This effect is restricted to the ENST00000452357 transcript, corroborating our recent finding that coExQTL are frequently isoform-specific^9^. Interestingly, this coExQTL effect at *OAS1* is not observed at the level of cytokine transcription, highlighting the utility of investigating regulatory networks across RNA and protein expression, but is also potentially in keeping with a non-canonical role for OAS1 isoforms in inflammatory responses, whereby *OAS1* isoforms support IRF1 translation on IFNγ stimulation^40^. The *OAS1* coExQTL that we identify here is a risk locus for severe COVID-19 disease^29^. Monocyte-derived hypercytokinaemia has been described as hallmark of severe COVID-19^41^, which taken alongside our data, suggests the possibility that *OAS1* may modify risk of severe disease through central regulation of cytokine production in activated monocytes.

Our study has a number of limitations. The number of cytokines that we were able to robustly quantify in this experiment was modest, and it is very likely that additional, disease-relevant insights would be delivered through expanding the number of analytes assayed. Further, gene expression was quantified at only two timepoints for LPS and one for IFNG using microarrays, compromising power to detect relationships between alternative splicing and cytokine secretion. Finally, while these data provide considerable insights into the regulation of cytokine production and its role in human disease, the models we use here reflect immune responses in healthy, European-ancestry adults. It will be important to expand our observations to understand the relevance of these findings across other populations, ages and health states.

In summary, by integrating genetic, RNA and protein expression data, we have comprehensively characterised the determinants of cytokine secretion in monocytes following innate immune stimulation. Our findings are highly informative of human disease risk and, in keeping with our previous work, highlight the importance of biological context in the design of functional genomic experiments to understand the determinants of human health.

## Materials & Methods

### Participants and biological sample processing

As previously^7^, monocytes were isolated from 432 healthy, European-ancestry adults aged 18-66 years (median age 30 years, 240 of which were female). Following venesection into EDTA-containing tubes (Vacutainer system, Becton Dickinson), PBMC separation was performed by density gradient centrifugation (Ficoll-Paque), monocytes were isolated by positive selection using CD14^+^ magnetic isolation kits (miltenyi). Monocytes were cultured at 500,000 cells/ml in RPMI with 5% fetal calf serum, penicillin/streptomycin and L-glutamine. Cultured monocytes from 261 and 322 donors were stimulated with 20ng/ml of LPS (Ultrapure LPS, Invivogen) for 2 and 24 hours respectively, and monocytes from 367 donors were stimulated for 24 hours with IFNγ (R&D Systems) at a final concentration of 20ng/ml. Monocytes from 77 donors were left unstimulated and cultured for 24 hours to act as untreated incubator controls. Following incubation, culture supernatants were harvested and stored at −80°C prior to subsequent cytokine quantification and monocytes were harvested and total RNA extracted (RNeasy Mini Kit, Qiagen). In addition, for each donor, genomic DNA was extracted from whole blood using Gentra Puregene Blood kits.

### Genotyping and imputation

Genome-wide genotyping was performed using with Illumina HumanOmniExpress-12v1.0 arrays. Following quality control, excluding SNPs with minor allele frequencies <1%, evidence of departure from Hardy Weinberg equilibrium (*P*<1×10^−6^) or call rates <98%, genotypes at 437,390 SNPs were taken forward for genome-wide imputation. Following exclusion of relatedness, samples were excluded if they were population (principal component) or heterozygosity outliers. Phasing and genotype imputation was performed with Eagle2^42^ and minimac^43^, using the Haplotype Reference Consortium (release 1.1) as a reference panel^44^, as implemented in the Michigan Imputation Server^45^. Following imputation, we excluded imputed SNPs with minor allele frequencies <5%, imputation quality (r^2^) <0.7 and evidence for departure from Hardy Weinberg equilibrium *P*<1×10^−10^. The resulted in 5,366,727 autosomal SNPs being included in downstream analysis. Calculation of QC metrics and sample/SNP exclusion was performed in QCTOOL (https://www.well.ox.ac.uk/~gav/qctool_v2/index.html) and PLINK^46^.

### Gene expression quantification

As previously^7^, gene expression was quantified using Illumina HumanHT-12 v4 BeadChip gene expression arrays, which includes 47,231 probes. Probes were excluded from downstream analysis if they mapped to more than one genomic region or a region with common genetic variation (minor allele frequency >1%). After quality control, 29,001 probes mapping to 18,078 genes were taken forward for downstream processing. Probe intensities were then normalised and variance stabilised (lumi), with batch correction using the ComBat package in R.

### Cytokine quantification

Protein concentrations of human cytokines, chemokines and growth factors were quantified in monocyte culture supernatants using multiplexed Luminex assays at the Thermo-Fisher laboratories (Bender MedSystems GmbH, Vienna, Austria). Protein quantification was performed using 45-plex Human Cytokine/Chemokine/Growth Factor Panel 1 ProcartaPlex kits (EPX450-12171-901, Affymetrix / Thermo-Fisher, Massachusetts, USA), customised with 5 additional analytes: CD-40, CD-40L, IDO, leptin and PD-L2. Assays were performed on a Luminex 200 machine according to manufacturer’s instructions and Affymetrix control samples. Samples were randomised with respect to treatment condition across 96-well plates and laboratory staff were blinded to sample treatment.

We quantified protein concentrations in monocyte culture supernatants in 1,059 samples from 366 individuals in 4 stimulation conditions: untreated, n=83; 2 hours LPS, n=278; 24 hours LPS, n=333; 24 hours IFNγ, n=364. We excluded 10 cytokines from downstream analysis as they were not detected above the lower limit of quantification in ≥10% of samples in ≥1 condition. In addition, we assayed 36 samples in duplicate (untreated, n=8; 2 hours LPS, n=8; 24 hours LPS, n=10; 24 hours IFNγ, n=10), excluding a further 12 cytokines from downstream analysis where the concordance between replicates was poor (Pearson’s correlation *P*>0.001, Fig. S7). We further excluded samples with low call rates, which we defined as failure to detect more than 2 cytokines above the lower limit of quantification for cytokines detected in ≥95% of samples in a given stimulation condition. Samples were also excluded where paired covariate, genotyping or gene expression data was incomplete.

Cytokine concentrations below the lower limit of quantification were assigned values of zero. Background (negative control sample) subtracted cytokines concentrations were normalised by inverse normal transformation, within stimulation condition and batch for analysis within a stimulation condition, and across all samples for analysis comparing conditions. Following normalisation, cytokine quantifications were residualised in a linear model to correct for age, sex and monocyte count, as well as the first five principal components of genome-wide genotyping data to correct for population structure in downstream genetic association analysis.

### Analysis of cytokine secretion

To define cytokine secretion induced by each stimulation condition, we compared normalised, covariate-corrected cytokine data between each stimulation condition (LPS 2 hours, LPS 24 hours and IFNγ 24 hours) with untreated incubator controls (cultured for 24 hours) in linear regression models. In addition, for each assayed cytokine, we extracted normalised transcriptomic data and compared stimulated gene expression in each condition with the untreated state in the same manner. To control for false discovery, we corrected p-values for the number of cytokines tested using the Benjamini-Hochberg method. We considered FDR<0.05 and an absolute log fold-change >2 to represent a significant change in cytokine secretion between conditions. Clustering of secreted cytokines in each condition was performed by hierarchical clustering of normalised, covariate-corrected cytokine concentrations, with 1 minus the absolute pairwise Pearson’s correlation coefficient used as the dissimilarity metric using the hclust() command in R.

To understand the degree to which RNA transcription of a cytokine is predictive of its secretion, we fitted linear regression models of paired, normalised RNA expression and protein secretion for each cytokine within each stimulation condition. We used model r^2^ to compare the predictive abilities of RNA expression and considered FDR<0.05 to be significant.

To test the effect of global baseline and stimulated transcriptional state on cytokine secretion we correlated baseline gene expression for each gene genome-wide with secretion of each cytokine in each stimulation condition, and further correlated RNA expression in the stimulated state with cytokine secretion. Analysis in each case was performed with linear regression, with the RNA gene expression matrix corrected for the first 20 principal components of gene expression. We applied two levels of false discovery control for this analysis, we first used the Benjamini-Hochberg method to correct for the number of genes tested, before applying a further Bonferroni correction to account for the number of cytokines tested. We considered an FDR<0.05 to be significant.

### Genome-wide association analysis

To define genetic predictors of cytokine secretion we performed multivariate and univariate genome-wide association analysis in each of the 3 stimulation conditions: LPS 2 hours, LPS 24 hours and IFNγ 24 hours. In each case we use normalised, covariate-corrected cytokine data, including correction for the first 5 principal components of genome-wide genotyping data to account for population structure. To account for imputation uncertainty, we used genotype dosages as predictors assuming an additive model of association. Multivariate association analysis was performed for the 28 cytokines passing quality control using multivariate analysis of variance (MANOVA) in R, using Pillai’s trace to derive an approximate F statistics. Univariate association testing was performed for each cytokine individually using linear regression in R.

### eQTL and co-expression QTL mapping

To define the eQTL for co-expression QTL mapping, we first mapped eQTL in each monocyte stimulation condition using the HRC-imputed genotyping data to ensure consistency of mapping across the gene expression and cytokine secretion data. We performed eQTL mapping for each gene in *cis*, correlating genotype dosage at SNPs within 1Mb of the transcription start site with normalised gene expression. Mapping was performed using QTLtools^47^, assuming an additive effect of SNP genotype on gene expression and including age, sex and the first 20 principal components of gene expression data. QTLtools approximates a permutation test to control false discovery at the level of the number of SNPs tested in *cis*, for which we performed 10,000 permutations. We then applied a second level of control for false discovery, controlling for the number of genes tested using the qvalue package in R.

No array probes for *CCL4* or *CCL5* expression passed quality control thresholds. To test the effect of the CCR5-Δ32 on transcription we therefore used genotyping and RNA-seq data from an analogous eQTL mapping experiment in naïve and stimulated stimulated monocytes^9^. These data represent an independent set of 185 healthy, European ancestry donors. Monocytes were isolated and cultured as described above, and cells were cultured for 24 hours without treatment or with LPS and IFNγ for 24 hours. RNA was quantified with paired-end 100bp sequencing on Illumina Hiseq-4000 machines, with read alignment using HISAT2^48^ and gene expression quantification with HTSeq^49^. Genome-wide genotyping was performed with Affymetrix UK Biobank Axiom arrays, with imputation of unassayed variants using SHAPEIT2/IMPUTE2^50,51^ with 1000 Genomes Project Phase 3 as a reference panel^44^. We extracted normalised *CCL4* and *CCL5* read counts and correlated these with genotype at rs113010081 (proxy for CCR5-Δ32) in a linear regression model corrected for age, sex and the first 20 principal components of the gene expression matrix. Post quality control, we correlated *CCL4*/*CCL5* gene expression with rs113010081 genotype in 176 (naïve monocytes), 176 (LPS-treated monocytes) and 139 (IFNγ-treated monocytes) individuals.

For co-expression QTL identification, we took peak eSNPs for high-confidence (FDR<1×10-5) *cis* eQTLs and restricted our analysis to peak eSNPs with a minor allele count of at least 5. We fitted linear regression models of eGene RNA expression as the dependent variable with cytokine secretion, eSNP genotype and an interaction term between eSNP genotype and cytokine secretion as predictors. We performed this analysis in each of the 3 stimulation conditions. We calculated interaction *P*-values using likelihood ratio tests, correcting for number of tests using the Benjamini-Hochberg method. We defined a significant coExQTL as a having a significant interaction term (FDR<0.05).

### Enrichment and colocalization

To define 95% credible SNP sets for our multivariate GWAS associations we used test statistics from the univariate analysis for the cytokine most associated at each locus. Where univariate GWAS identified experiment-wide significant association we used these test statistics directly. For each locus, we took SNPs within 100kb of the peak association and computed approximate Bayes’ factors from betas and standard errors^52^, assuming a prior distribution of N(0; 0.2^2^). All SNPs in the window were considered to have an equal prior probability of being the causal SNP. The 95% credible SNP set was defined as the fewest SNPs whose summed posterior probabilities exceed 95%.

To define the likelihood that our cytokine GWAS signals share a causal variant with human GWAS loci we used a Bayesian colocalization approach implemented in Coloc v5.1.0^53^. We assessed evidence for colocalization among summary statistics from 47 UK biobank traits (https://www.nealelab.is/uk-biobank/, Table S23) and 100 GWAS Catalog traits (Table S24), testing for colocalization where there is evidence of genome-wide significant association (*P*<5×10^−8^) within a 200kb window centred on the lead cytokine GWAS SNP. To understand disease/trait informativeness of coExQTL we used the CausalDB resource^54^ to understand whether our lead SNP is within 95% credible sets of published GWAS. Throughout we tested for enrichment of genes of interest, for instance component genes of transcriptional networks associated with cytokine secretion, against a background of all genes tested using hypergeometric tests implemented in XGR^55^. We used GOBP and Disease Ontology gene lists as query ontologies, as well as single cell transcriptome data defining cell-type specific transcripts to identify platelet-specific genes^56^. We corrected enrichment *P*-values for the number of terms tested using the Benjamini-Hochberg method.

## Data Availability

Gene expression data are available through ArrayExpress (accession: E-MTAB-2232). Genotyping data have been deposited at the European Genome-phenome Archive and are available on request (EGAS00000000109). Cytokine measurements will be made available on publication. All data produced in the present study are available upon reasonable request to the authors.

## Acknowledgements

We thank Francesca Liu and Markus Miholts from Thermo-Fisher for their help planning, performing and analysing the ProcartaPlex assay. JJG and AJM were supported by NIHR Academic Clinical Lectureships. The work was supported by a Wellcome Trust Career Development Award (311117/Z/24/Z) to JJG, a Wellcome Career Development Award (226535/Z/22/Z) to BPF, a Wellcome grant (098759/Z/12/Z) to LJ and the Newton Fund (FONCICYT/50/2016). JCK was supported by a Wellcome Trust Investigator Award (204969/Z/16/Z), the National Institute for Health Research (NIHR) Oxford Biomedical Research Centre and the Chinese Academy of Medical Sciences (CAMS) Innovation Fund for Medical Science (grant 2018-I2M-2-002). The research was supported by the Wellcome Trust Core Award Grant Number 203141/Z/16/Z with additional support from the NIHR Oxford BRC. The views expressed are those of the author(s) and not necessarily those of the NHS, the NIHR or the Department of Health.

