## Supplementary Figures for "Genetic determinants of cytokine production in activated human monocytes"

### Supplementary Data Table Legends

**Supplementary Table 1.** Changes in monocyte cytokine secretion induced by innate immune stimulation. Fold-change and p-values are calculated with respect to cytokine secretion from monocytes without stimulation (n=78). FDR, false discovery rate.

**Supplementary Table 2.** Correlation between cytokine RNA expression and protein secretion in monocytes stimulated with LPS and IFN$\gamma$. FDR, false discovery rate.

**Supplementary Table 3.** Prediction of cytokine secretion from baseline transcriptional state in LPS and IFN$\gamma$-stimulated monocytes. FDR, false discovery rate.

**Supplementary Table 4.** Prediction of cytokine secretion from the stimulated transcriptional state in LPS and IFN$\gamma$-stimulated monocytes. Where ≥10 genes are associated with secretion of a cytokine, we performed enrichment analysis using the REACTOME database. FDR, false discovery rate.

**Supplementary table 5.** Genomic inflation factors for multivariate and univariate genome-wide association analyses in stimulated monocytes.

**Supplementary table 6.** Effect of rs5757584:T allele carriage on cytokine secretion in stimulated monocytes.

**Supplementary table 7.** Transcription factor binding sites overlapping with rs5757584.

**Supplementary table 8.** Evidence for colocalization between the genetic signal for PDGF-BB secretion in IFN$\gamma$-stimulated monocytes at rs5757584 and GWAS signals of human phenotypic traits (UK biobank) and disease risk (GWAS Catalog). GWAS are included if there is a significant locus ($P<5x10^{-8}$) within 100kb of rs5757584.

**Supplementary table 9.** Effect of rs11123160:T allele carriage on cytokine secretion in stimulated monocytes.

**Supplementary Supp 10.** Transcription factor binding sites overlapping with rs11123160.

**Supplementary table 11.** Evidence for colocalization between the genetic signal for IL-1RA secretion in LPS-stimulated (24-hour) monocytes at rs11123160 and GWAS signals of human phenotypic traits (UK biobank) and disease risk (GWAS Catalog).

GWAS are included if there is a significant locus ($P<5x10^{-8}$) within 100kb of rs11123160.

**Supplementary table 12.** Effect of rs113010081:C allele carriage on cytokine secretion in stimulated monocytes.

**Supplementary table 13.** Evidence for colocalization between the genetic signal for MIP-1β secretion in IFNγ-stimulated monocytes at rs113010081 and GWAS signals of human phenotypic traits (UK biobank) and disease risk (GWAS Catalog). GWAS are included if there is a significant locus ($P<5x10^{-8}$) within 100kb of rs113010081.

**Supplementary table 14.** Effect of rs13296842:G allele carriage on cytokine secretion in stimulated monocytes.

**Supplementary Table 15.** Lead eSNP summary statistics for high-confidence eQTLs ($FDR<1x10^{-5}$) in 2-hour LPS-stimulated monocytes.

**Supplementary Table 16.** Lead eSNP summary statistics for high-confidence eQTLs ($FDR<1x10^{-5}$) in 24-hour LPS-stimulated monocytes.

**Supplementary Table 17.** Lead eSNP summary statistics for high-confidence eQTLs ($FDR<1x10^{-5}$) in 24-hour IFNγ-stimulated monocytes.

**Supplementary Table 18.** Significant RNA:cytokine secretion coexpression QTLs in 2-hour LPS-stimulated monocytes.

**Supplementary Table 19.** Significant RNA:cytokine secretion coexpression QTLs in 24-hour LPS-stimulated monocytes.

**Supplementary Table 20.** Significant RNA:cytokine secretion coexpression QTLs in 24-hour IFNγ-stimulated monocytes.

**Supplementary Table 21.** Significantly enriched Gene Ontology Biological Process terms among genes with RNA:cytokine secretion coexpression QTLs.

**Supplementary Table 22.** Significantly enriched Disease Ontology terms among genes with RNA:cytokine secretion coexpression QTLs.

**Supplementary Table 23.** Details of UK Biobank GWAS traits (n=47) used in colocalization analysis.

**Supplementary Table 24.** Details of GWAS Catalog GWAS traits (n=100) used in colocalization analysis.

### Supplementary Figures

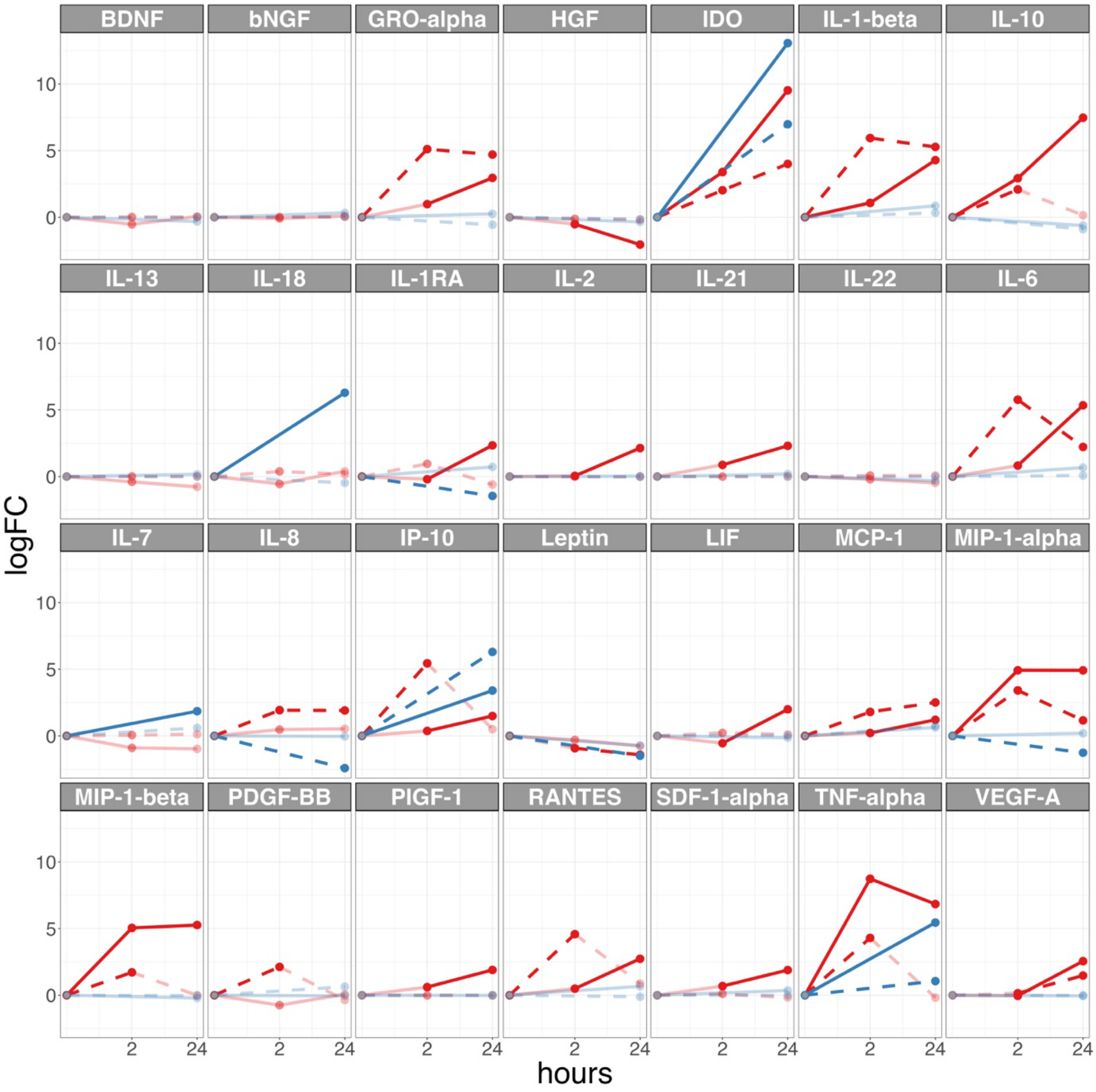


**Figure S1. Protein and transcriptional cytokine responses to innate immune stimulation.** Red lines represent responses to LPS, blue lines responses to IFNγ. Solid lines represent cytokine secretion, dashed lines represent RNA expression. Lines in bold represent significant changes (FDR<0.05, fold change ≥2).

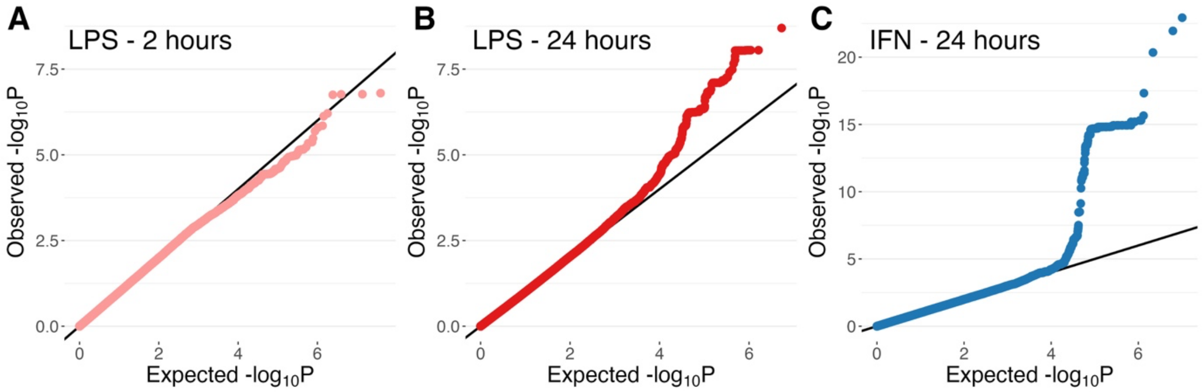


**Figure S2. QQ plots for genome-wide multivariate association analysis.** Genome-wide multivariate association analysis MANOVA of secretion of 28 cytokines from monocytes stimulated with LPS (2 hours, pink, panel A), LPS (24 hours, red, panel B) and IFNγ (24 hours, blue, panel C).

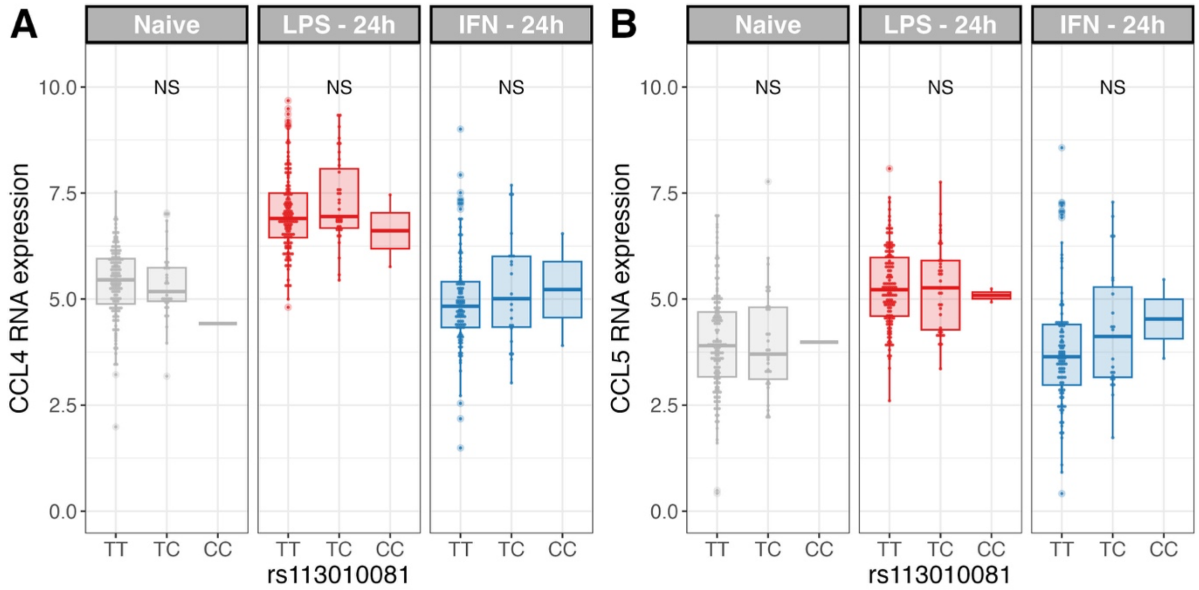


**Figure S3. Effect of CCR5-Δ32 on *CCL4*/*CCL5* RNA transcription.** (**A**) Effect of rs113010081 genotype (proxy for CCR5-Δ32) on *CCL4* RNA expression in naïve monocytes and monocytes stimulated for 24 hours with LPS (red) and IFNγ (blue). (**B**) Effect of rs113010081 genotype (proxy for CCR5-Δ32) on *CCL5* RNA expression in naïve monocytes and monocytes stimulated for 24 hours with LPS (red) and IFNγ (blue). Box and whisker plots; boxes depict the upper and lower quartiles of the data, and whiskers depict the range of the data excluding outliers (outliers are defined as data-points >1.5X the inter-quartile range from the upper or lower quartiles).

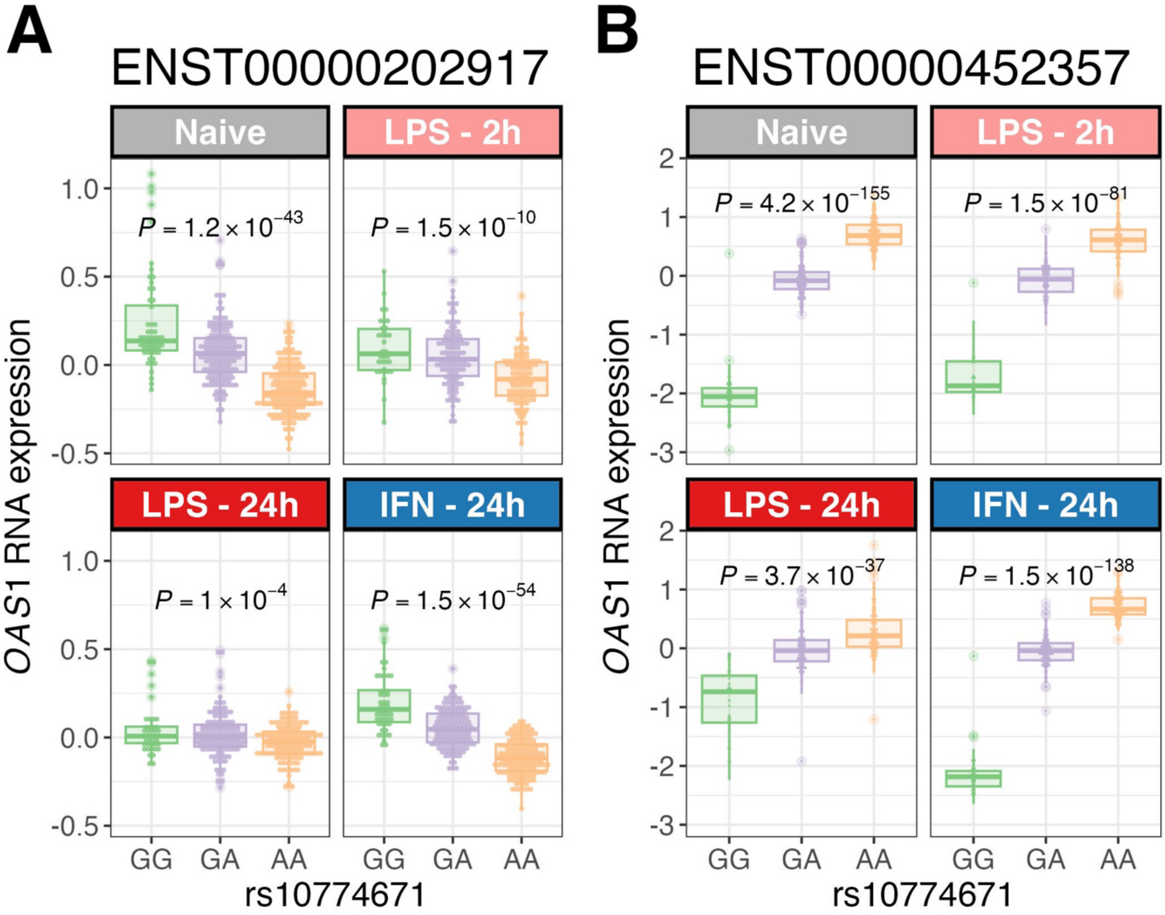


**Figure S4. Effect of rs10774671 on *OAS1* isoform usage.** (**A**) Effect of rs10774671 genotype ENST00000202917 OAS1 transcript expression naïve monocytes and stimulated monocytes. (**B**) Effect of rs10774671 genotype ENST00000452357 OAS1 transcript expression naïve monocytes and stimulated monocytes. Box and whisker plots; boxes depict the upper and lower quartiles of the data, and whiskers depict the range of the data excluding outliers (outliers are defined as data-points >1.5X the inter-quartile range from the upper or lower quartiles).

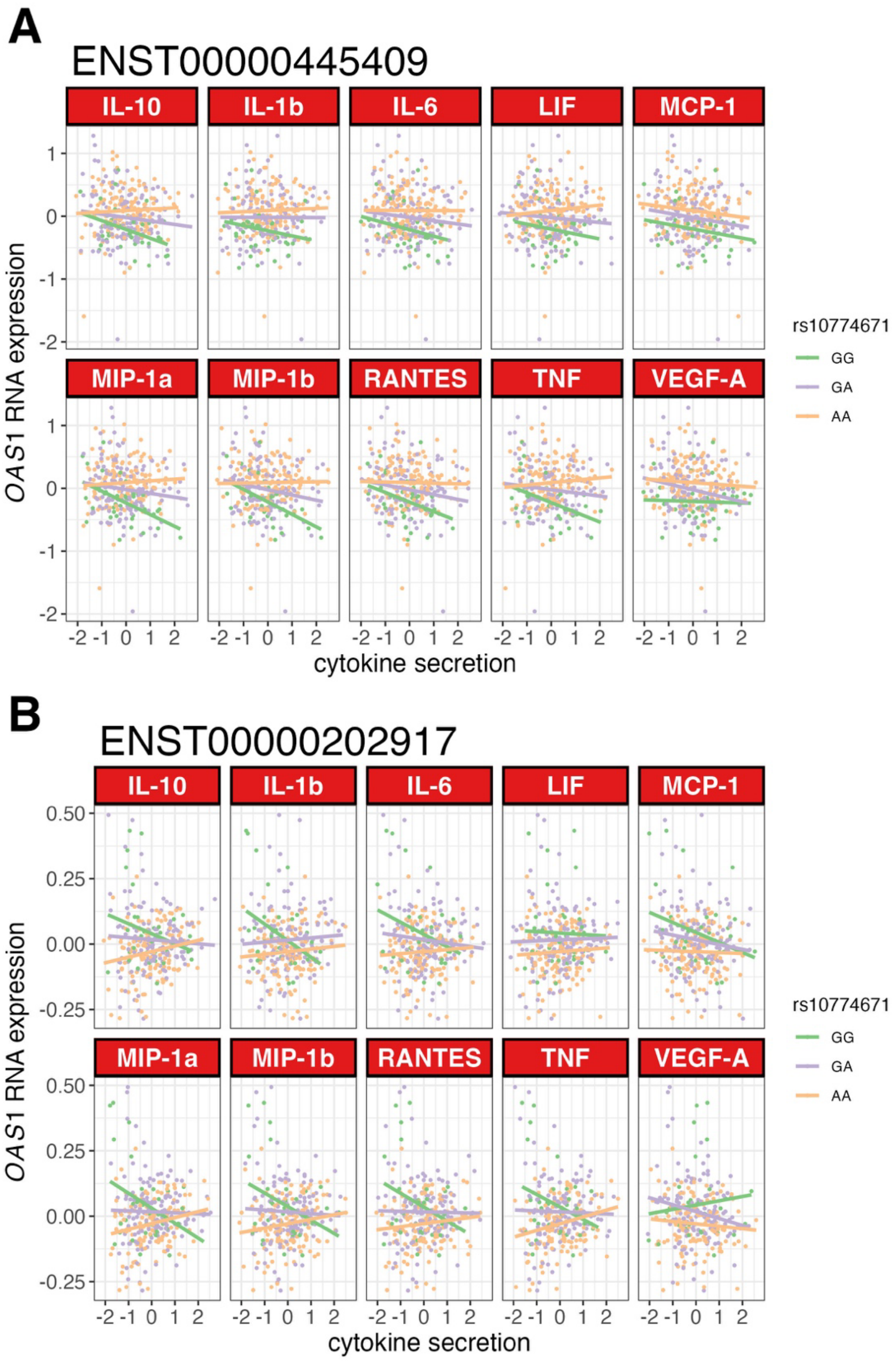


**Figure S5. Evidence for rs10774671 acting as co-expression QTL with alternative *OAS1* isoforms.** Evidence of a coExQTL effect for rs10774671 genotype on the relationship between 10 *OAS1* coExQTL-associated cytokines and expression of the canonical OAS1 transcript, ENST00000445409 (**A**) and ENST00000202917 (**B**). Tests for interaction with either transcript are null (FDR>0.05) for all transcript:cytokine pairs.

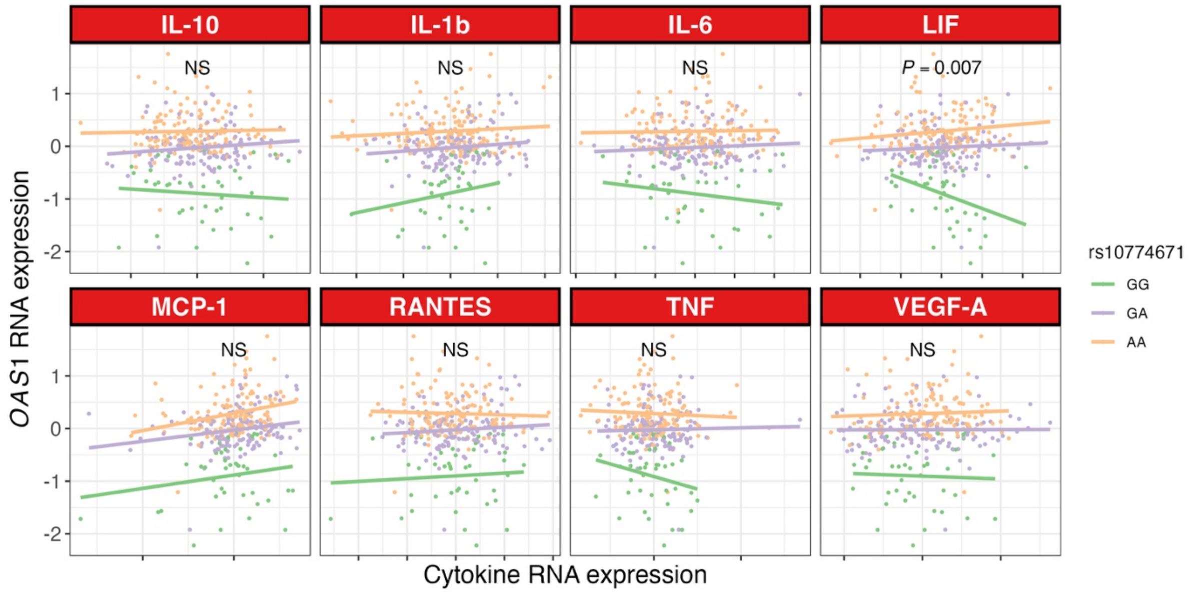


**Figure S6. Evidence for rs10774671 acting as co-expression QTL between *OAS1* cytokine RNA expression.** Evidence of coExQTL effects for rs10774671 genotype on the relationship between 8 *OAS1* coExQTL-associated cytokines and expression of OAS1 (ENST00000452357). P-values represent test of interaction. NS, not significant.

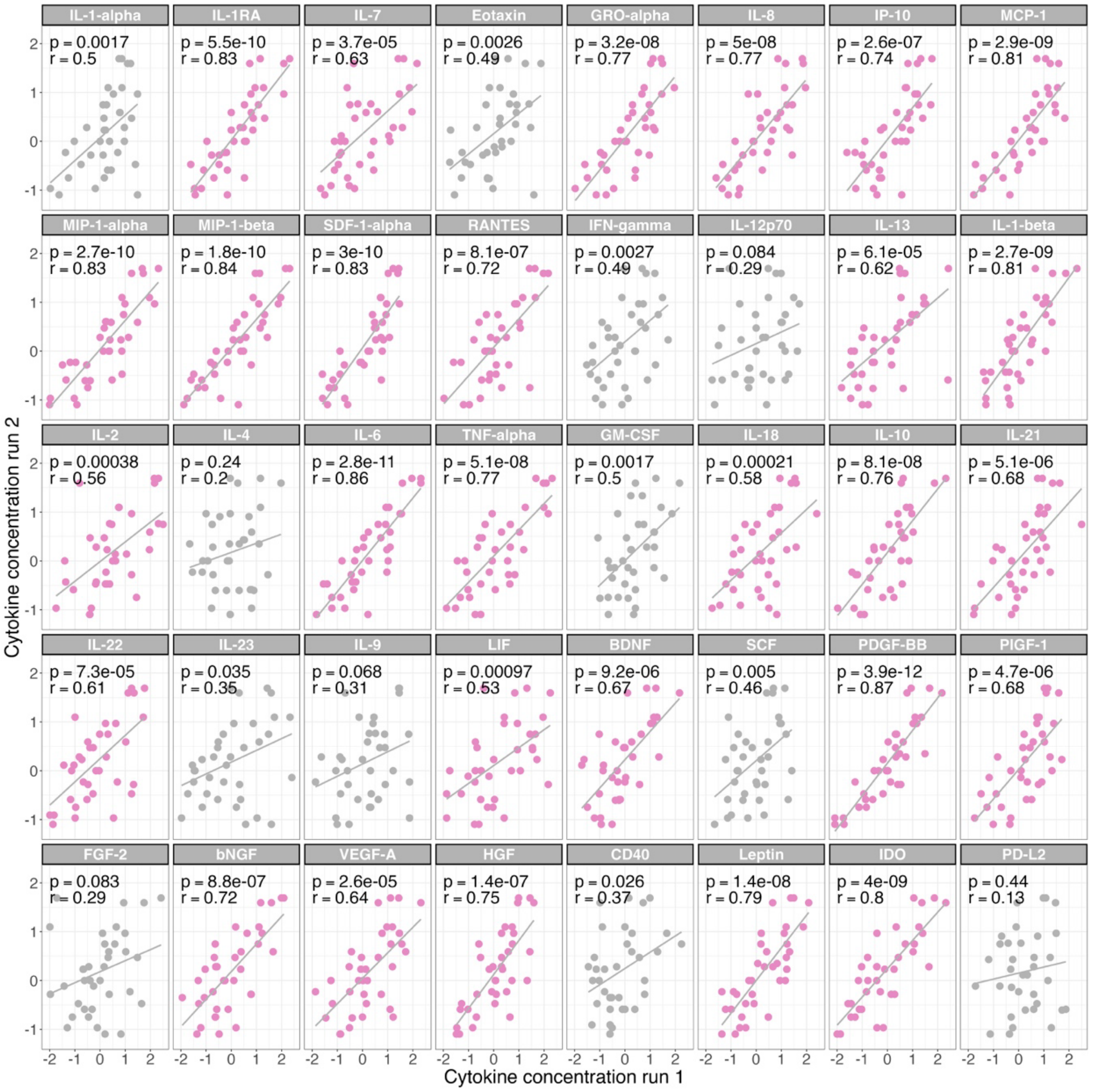


**Figure S7. Cytokine quantification quality controls.** Cytokine quantification estimates for 50 cytokines in 36 samples assayed in duplicate. Agreement between assay replicates is calculated by Pearson's correlation with normalised data. Cytokines passing the quality control threshold (*P*<0.001) are highlighted in pink.
